# The Effect of Psychotherapy in the Management of Obesity: An umbrella review

**DOI:** 10.1101/2024.10.19.24315815

**Authors:** Mariana Vasquez-Ponce, Óscar Gómez, Luisa F. González Ballesteros, Dina Peralta-Reich, Julieta Bermúdez-Gómez, Juan David Junco, Maria Alejandra Alba, Luisa Suarez-Ordoñez, Nayeh Arana, Eduardo Ballesteros, Valeria Mojica-Sierra, Lina María González Ballesteros

## Abstract

**Objective:** To assess the impact of psychotherapy on obesity treatment in outpatient settings.

**Methods:** The study targets adults with a BMI of 30 or higher, comparing psychotherapeutic interventions to usual care or no treatment. Key outcomes include weight loss, adverse events, depression, quality of life, and long-term BMI changes. Extensive database searches were conducted.

**Results:** Out of 1,334 articles reviewed, only 3 studies met the inclusion criteria, emphasizing the scarcity of relevant research. These studies primarily focused on behavioral and cognitive-behavioral strategies. Results showed that combining behavior therapy with diet/exercise led to greater weight loss than diet/exercise alone. Studies on cognitive therapy and general psychotherapy were less frequent and yielded weaker evidence. Despite varied methodologies, the findings suggest that behavioral therapy combined with dietary and exercise interventions offers improved outcomes, though more research is needed to strengthen these conclusions.

**Conclusions:** The study found that combining behavioral therapy with diet and exercise resulted in greater weight loss than diet and exercise alone. However, research on the impact of psychotherapy on obesity is limited, as only 3 studies met the inclusion criteria. Cognitive therapies and general psychotherapy provided weaker evidence. Therefore, further research is necessary to validate these findings and investigate alternative psychotherapeutic methods for treating obesity.

## 1. INTRODUCTION

Obesity is a complex disease defined as having a body mass index (BMI) ≥30 kg/m^2^ by excessive adipose tissue accumulation along with systemic inflammation, resulting from an energy imbalance where caloric intake surpasses expenditure (1). Genetic predisposition, metabolic and endocrine alterations, environmental factors, and lifestyle behaviors influence its pathogenesis (2-4). The presence of obesity has been associated with a higher risk of chronic, noncommunicable diseases and adverse health outcomes, such as cardiovascular disease, diabetes, osteoarthritis, hyperlipidemias and recently, with some types of cancer (5). It poses a global public health issue, as excessive weight gain has significantly increased since 1980. This trend is seen in various countries, both males and females and in all age groups (5-7).

Additionally, obesity treatments, including pharmacological and surgical methods, have mixed results and concerns about long-term effects. Health policies like taxes and advertising restrictions have been effective in the short term (8). However, behavioral and biological factors limit long-term treatment planning (9).

Psychotherapy, as defined by the American Psychiatric Association (APA), is a treatment for health conditions and emotional challenges, aiming to alleviate symptoms and identify root causes. Families or groups can apply it, with weekly sessions lasting 45–50 minutes, depending on patient goals and frequency (10). The APA reports that 75% of participants have reported benefits from the program, with the success influenced by the type of psychotherapy used (8). Although there are no adverse or side effects reported in this matter. A pertinent observation highlights the need for further research to establish comprehensive theoretical definitions, classifications, and evaluations of these side effects, as they remain insufficiently addressed in the current literature (11).

Obesity is caused by a combination of physiological, genetic, psychosocial, and environmental factors. Psychological interventions are crucial in managing obesity, despite being often overshadowed by medical approaches (12-15). Studies show a link between behavioral factors like diet and sedentary lifestyle with obesity (16). Behavioral therapy, along with dietary and exercise interventions, is recommended for overweight individuals. Cognitive and behavioral interventions aim to change negative habits and promote positive lifestyles, and constitute the main interventions from this perspective (17-19).

There is an increasing number of systematic reviews on behavioral interventions for obesity, with recent research suggesting that comprehensive and intensive interventions can lead to modest weight loss in severely patients with obesity (20-22), nevertheless, heterogeneity of the results is mainly attributed to differences in population and the available therapeutic approaches (8). Interest on this topic varies from analyzing the best approach to investigating the effect of combining different therapies. To address gaps in research, a holistic overview including psychotherapeutic approaches other than behavior therapy are needed. An umbrella review is essential for an integrated analysis of existing systematic reviews on this topic.

## 2. METHODS

The umbrella review was done by the research team following a set of rules, and it was reported using the PRISMA (23) guidelines for preferred reporting items for systematic reviews and meta-analyses. The protocol is available in the supplementary material.

### 2.1 Eligibility criteria

Studies were included if they met the following criteria: 1) systematic review of randomized clinical trials on the treatment of obesity (defined as BMI≥ 30 kg/m2 or above the 95th percentile in children/adolescents) with a known form of psychotherapy; 2) participants aged 8 to 65 years; 3) the primary outcome was weight loss assessed in at least one follow-up; 4) published in English or Spanish. Systematic reviews were excluded if they included samples with eating disorders, bariatric surgery, or secondary forms of obesity, were focused on obesity prevention, or described the intervention as behavioral therapy.

### 2.2. Information sources

Eligible articles were identified by searching MEDLINE (PubMed), EMBASE (EMBASE), Cochrane Database of Systematic Reviews (Ovid), PsycArticles (PsycNet), Scopus (Scopus), and ProQuest Dissertations and Theses (ProQuest), from inception through March 4th, 2024. All studies were written in English and published between 2001 and 2024. The search strategy developed for the PubMed interface is detailed in the supplementary material. This strategy was adapted to the appropriate syntax and applied to the other included databases. Additionally, we reviewed the reference lists of retrieved systematic reviews for any relevant publications.

### 2.3 Selection process and Data extraction

Four teams of reviewers (MV and OG, NA and JJ, MA and JB, or VM and LG), independently screened all records to assess eligibility using Rayyan systematic review software (24). The eligible reviews were retrieved and evaluated in full text by two authors separately, with any disagreements resolved by consensus of the entire team. The agreement between reviewers was 96%. Subsequently, teams of two reviewers independently extracted data from each systematic review into an Excel spreadsheet. For each review, we collected information on the inclusion and exclusion criteria, the number and design of included studies, the country, the types of psychotherapies, the effect measures, the quality assessment, and whether meta-analyses were conducted. Each team compared the independently extracted data, and disagreements were resolved by a senior reviewer.

### 2.4 Bias assessment of included studies

Two reviewers independently used the AMSTAR 2 tool (25) to assess the quality of the included systematic reviews, with any disagreements discussed and resolved by consensus. Briefly, the AMSTAR 2 qualifies the confidence in a systematic review result in one of four groups, ranging from critically low to high.

### 2.5 Data Analysis

In the protocol of this umbrella review, we aimed to analyze the effects of psychotherapy using a random-effects meta-analytic approach when sufficient studies were included and the systematic reviews reported outcomes as effect sizes that could be combined in the analysis. However, as described below, these conditions were not met, and the included studies were described qualitatively. Additionally, a modification in the exclusion criteria was necessary because all the reports included in the review involved overweight patients. This adaptation was implemented to ensure the consistency and comparability of the results.

## 3. RESULTS

A total of 1334 articles were identified; after the exclusion of duplicates, 1092 articles remained, of which, only 78 articles were retrieved for full-text examination. 3 systematic reviews met inclusion criteria, as shown in Figure 1.

Table 1 provides a detailed description of the selected articles. Overall, the main interventions studied included CBT, behavior therapy, mindfulness, family-based interventions, and family therapy combined with other interventions such as calorie intake reduction strategies and psychoeducation. The results were found to be described in terms of BMI and weight change. Still, there was considerable difficulty in the interpretation of results because of the heterogeneity related to the combination of interventions described as “psychotherapy” and the lack of consensus in the description of the measure units for the diagnosis. After adjusting the exclusion criteria to include only studies specifying overweight patients, the total analyzed sample met the new parameters, allowing for a more precise evaluation of the intervention effects in this specific population.

Three systematic reviews evaluated different obesity interventions, analyzing a total of 79 randomized controlled trials (RCTs). The article by Sung-Chang et al. looked at 15 RCTs on family-based interventions. It gave each study an average methodological score of 7.93 out of 14, which means that 10 of them used rigorous and well-aligned methods. Interventions combining family therapy, lifestyle changes, and parent education showed the greatest effect, although methodological heterogeneity made it difficult to analyze specific outcomes. Han Shi et al., on the other hand, looked at 33 RCTs that compared cognitive-behavioral therapy (CBT) and behavioral therapy. They found that both helped people lose weight, though two studies didn’t find any significant changes in BMI. Finally, Han Shi et al. looked at 31 RCTs to see how well weight-loss programs that target emotional eating worked. They found that these programs had small to moderate effects on binge eating but no significant effects on BMI right after the program. There was no conclusive evidence on the effectiveness of combining these interventions with other therapeutic strategies.

In these studies, the assessment of bias was approached in different ways, the article of Sung-Chang et al. was assessed only by the selection of RCTs in the study. On the other hand, Han Shi et al. based on the funnel plot and use of the Cochranes risk of bias tool found no significant publication bias in their sample, as did Han Shi et al. which also assessed through the Eggers test, finding no publication bias.

### 3.1 Primary outcome

Weight loss assessed through changes in BMI depended on the psychotherapeutic intervention analyzed. Regarding cognitive behavioral therapy as an intervention, Chew et al. found that BMI reduction was more effective in studies that included a control group, with an average decrease of 3–5% compared to studies without a control group. However, this difference in BMI reduction only became significant 12 months post-intervention. On the contrary, Mesaric K. et al. obtained a moderately significant short-term effect in some of the studies included in their report. However, they noted follow-up and adherence limitations. Family therapy showed better results when combined with parent education and lifestyle interventions.

### 3.2 Secondary outcomes

The secondary outcomes primarily pertain to quality of life, specifically enhancing emotional eating. A small to medium effect was observed with CBT; however, these results were only significant during the first 3 months post-intervention. Similarly, a small to medium effect was noted in retraining eating habits, with no significant effect on binge eating.

## 4. DISCUSSION

Obesity currently affects over 40% of adults globally, impacting over 2.6 billion people’s health, quality of life, and life expectancy. Additionally, around 1.3 billion adults are living with overweight. In many parts of the world, access to bariatric surgery, considered the gold standard for treating severe obesity, is limited. Consequently, most individuals with obesity must seek nonsurgical weight loss options. Because of these problems, new patient-centered research has shown that we need to learn more about and prioritize effective nonsurgical weight loss interventions to make things better for people who can’t get surgery (26).

This comprehensive and up-to-date study investigated the effectiveness of various psychotherapy approaches in treating obesity in adults.

Our findings suggest that patient education and psychotherapy significantly improved BMI reduction despite previous reports indicating a small to medium impact. The effect was more pronounced when the time-lapse exceeded 12 months, highlighting the primary influence of psychotherapeutic interventions during the maintenance phases, a crucial aspect of traditional obesity treatments. There is a need to establish universal diagnostic criteria and definitions of psychotherapy in the current and/or future studies to advance in the standardization to establish the effectiveness of psychotherapy in the management of obesity as an adjunctive therapy to the usual approaches.

Identifying three major weaknesses: 1) no clear distinction between patients with overweight or obesity which reflects the divergence and absence of consensus in the diagnostic criteria. 2) issues with the intervention: as many reports lacked a clear definition of psychotherapy or did not specify the intervention, related to wide definitions of behavior therapy. 3) the outcomes were not aligned with our objective of analyzing changes in BMI. Furthermore, the majority of the discarded articles featured primary and secondary outcomes that addressed qualitative variables, lacking any standardization or heterogeneity analysis. Therefore, we considered it a weakness not to have objective outcomes to compare among different interventions, including behavioral change and psychotherapy interventions.

Modification of the exclusion criteria was necessary because all the reports included overweight patients. Initially, many articles were identified in each study, however, the sample size was drastically reduced when inclusion and exclusion criteria were applied. Despite the lack of clarity in most studies analyzed, regarding whether the intervention applied was “behavioral” or psychotherapy itself, we did not modify this inclusion criterion nor were we flexible about it, excluding any study that did not indicate that it involved strict psychotherapeutic intervention.

In conclusion, psychotherapy clearly impacts obesity due to its correlation with psychological variables. However, compared to behavioral interventions alone, psychotherapy has the benefit of maintaining the results.

### Strengths

This study has several notable strengths. The research question was shaped by input from both adults and children experiencing obesity, leading to a relevant and significant review with broadly applicable findings. We collaborated with medical experts (LG and DP) to create a thorough search strategy designed to identify studies pertinent to our research question. The review adhered to PRISMA reporting guidelines and employed the AMSTAR2 tool were employed to evaluate the quality of reporting in the included studies, thereby ensuring a rigorous and transparent presentation of methods.

### Limitations

Despite not having a formal registration, the study followed a predefined written protocol, adhered to established guidelines, and reported using PRISMA criteria. Another limitation is that, while we aimed to include studies with a minimum intervention duration of 12 months, some individual studies within the systematic reviews or meta-analyses had shorter durations. However, we were able to exclude these studies. Additionally, most of the included studies focused on females and children, indicating a selection bias that may limit the generalizability of the results to other populations, such as males.

### Future research

Establishing universal diagnostic criteria to distinguish between overweight and persons with obesity is essential to address the current divergence in the field. Additionally, studies should clearly define and standardize psychotherapeutic interventions, particularly in differentiating between behavioral therapy and other forms of psychotherapy. Using standardized measures to highlight objective outcomes, such as BMI changes, will allow for better comparison across interventions. Moreover, expanding research to include diverse populations, particularly Latin American populations, is critical to understanding regional variations and improving the generalizability of findings. Longer intervention durations should also be explored to better assess the long-term efficacy of psychotherapeutic approaches in obesity treatment.

## Supporting information

Appendix 1. Search strategy

Table 1. Main findings provided by the selected articles

Figure 1: Flow diagram of the identification, screening, and selection of studies for this Umbrella review.

## Data Availability

All data produced in the present work are contained in the manuscript

