## Appendix 1. Search strategy for "The Effect of Psychotherapy in the Management of Obesity: An umbrella review"

| | Query | Results |
|---|---|---|
| 1 | Obesity[MeSH Terms] | 268,588 |
| 2 | Obesity[Title/Abstract] | 336,461 |
| 3 | "adipose tissue hyperplasia"[Title/Abstract] OR adiposit*[Title/Abstract] OR "body weight"[Title/Abstract] OR "body weight excess"[Title/Abstract] OR "fat overload syndrome"[Title/Abstract] | 275,632 |
| 4 | ((Obesity[MeSH Terms]) OR (Obesity[Title/Abstract])) OR ("adipose tissue hyperplasia"[Title/Abstract] OR adiposit*[Title/Abstract] OR "body weight"[Title/Abstract] OR "body weight excess"[Title/Abstract] OR "fat overload syndrome"[Title/Abstract]) | 639,069 |
| 5 | Psychotherapy[MeSH Terms] | 222,126 |
| 6 | Psychotherapy[Title/Abstract] | 45,524 |
| 7 | Psychotherapies[Title/Abstract] | 2,63 |
| 8 | Psychotherapeutic[Title/Abstract] | 10,574 |
| 9 | (((Psychotherapy[MeSH Terms]) OR (Psychotherapy[Title/Abstract])) OR (Psychotherapies[Title/Abstract])) OR (Psychotherapeutic[Title/Abstract]) | 239,244 |
| 10 | ("Systematic Review"[Publication Type:NoExp] OR "Systematic Reviews as Topic"[mesh:noexp] OR (("comprehensive"[TIAB] OR "integrated"[TIAB] OR "integrative"[TIAB] OR "mapping"[TIAB] OR "methodology"[TIAB] OR "narrative"[TIAB] OR "scoping"[TIAB] OR "systematic"[TIAB]) AND ("search"[TIAB] OR "searched"[TIAB] OR "searches"[TIAB] OR "studies"[TIAB]) AND ("cinahl"[TIAB] OR "cochrane"[TIAB] OR "embase"[TIAB] OR "psycinfo"[TIAB] OR "pubmed"[TIAB] OR "medline"[TIAB] OR "scopus"[TIAB] OR "web science"[TIAB] OR "bibliographic review"[TIAB:~1] OR "bibliographic reviews"[TIAB:~1] OR "literature review"[TIAB:~1] OR "literature reviews"[TIAB:~1] OR "literature search"[TIAB:~1] OR "literature searches"[TIAB:~1] OR "narrative review"[TIAB:~1] OR "narrative reviews"[TIAB:~1] OR "qualitative review"[TIAB:~1] OR "qualitative reviews"[TIAB:~1] OR "quantitative review"[TIAB] OR "quantitative reviews"[TIAB])) OR "comprehensive review"[TIAB] OR "comprehensive reviews"[TIAB] OR "comprehensive search"[TIAB] OR "comprehensive searches"[TIAB] OR "critical review"[TIAB] OR "critical reviews" [TIAB] OR (("electronic database"[TIAB:~1] OR "electronic databases"[TIAB:~1] OR "databases searched"[TIAB:~3]) AND (eligibility[tiab] OR excluded[tiab] OR exclusion[tiab] OR included[tiab] OR inclusion[tiab])) OR "evidence assessment"[TIAB] OR "evidence review"[TIAB] OR | 521,863 |

| | | |
|---|---|---|
| | "exploratory review"[TIAB] OR "framework synthesis"[TIAB] OR "Integrated review"[TIAB] OR "integrated reviews"[TIAB] OR "integrative review"[TIAB:~1] OR "integrative reviews"[TIAB:~1] OR "mapping review"[TIAB:~1] OR "meta-review"[TIAB:~1] OR "meta-synthesis"[TIAB:~1] OR "methodology review"[TIAB:~1] OR ("mixed methods"[TIAB:~0] AND "methods review"[TIAB:~1]) OR ("mixed methods"[TIAB:~0] AND "methods synthesis"[TIAB:~1]) OR "overview reviews"[TIAB:~4] OR ("PRISMA"[TIAB] AND "preferred"[TIAB]) OR "PRISMA-P"[TIAB:~0] OR "prognostic review"[TIAB:~1] OR "psychometric review"[TIAB:~1] OR ("rapid evidence"[TIAB:~0] AND "evidence assessment"[TIAB:~0]) OR "rapid realist"[TIAB:~0] OR "rapid review"[TIAB:~1] OR "rapid reviews"[TIAB:~1] OR "realist review"[TIAB:~1] OR "review of reviews"[TIAB:~1] OR "scoping review"[TIAB:~1] OR "scoping reviews"[TIAB:~1] OR "scoping study"[TIAB:~1] OR ("state art "[TIAB:~2] AND "art review"[TIAB:~1]) OR "systematic evidence map"[TIAB] OR "systematic mapping"[TIAB:~1] OR "systematic literature"[TIAB:~1] OR "systematic Medline"[TIAB:~2] OR "systematic PubMed"[TIAB:~2] OR "systematic review"[TIAB:~1] OR "systematic reviews"[TIAB:~1] OR "systematic search"[TIAB:~1] OR "systematic searches"[TIAB:~1] OR "systematical review"[TIAB:~1] OR "systematical reviews"[TIAB:~1] OR "systematically identified"[TIAB:~1] OR "systematically review"[TIAB:~1] OR "systematically reviewed"[TIAB:~1] OR "umbrella review"[TIAB:~1] OR "umbrella reviews"[TIAB:~1] OR "Cochrane Database Syst Rev"[ta] OR "evid rep technol assess full rep"[Journal] OR "evid rep technol assess summ"[Journal]) | |
| 11 | ((((Obesity[MeSH Terms]) OR (Obesity[Title/Abstract])) OR ("adipose tissue hyperplasia"[Title/Abstract] OR adiposit*[Title/Abstract] OR "body weight"[Title/Abstract] OR "body weight excess"[Title/Abstract] OR "fat overload syndrome"[Title/Abstract])) AND ((((Psychotherapy[MeSH Terms]) OR (Psychotherapy[Title/Abstract])) OR (Psychotherapies[Title/Abstract])) OR (Psychotherapeutic[Title/Abstract]))) AND (("Systematic Review"[Publication Type:NoExp] OR "Systematic Reviews as Topic"[mesh:noexp] OR (("comprehensive"[TIAB] OR "integrated"[TIAB] OR "integrative"[TIAB] OR "mapping"[TIAB] OR "methodology"[TIAB] OR "narrative"[TIAB] OR "scoping"[TIAB] OR "systematic"[TIAB]) AND ("search"[TIAB] OR "searched"[TIAB] OR "searches"[TIAB] OR "studies"[TIAB]) AND ("cinahl"[TIAB] OR "cochrane"[TIAB] OR "embase"[TIAB] OR "psycinfo"[TIAB] OR "pubmed"[TIAB] OR "medline"[TIAB] OR "scopus"[TIAB] OR "web science"[TIAB] OR "bibliographic | 363 |

| | |
|---|---|
| | review"[TIAB:~1] OR "bibliographic reviews"[TIAB:~1] OR "literature review"[TIAB:~1] OR "literature reviews"[TIAB:~1] OR "literature search"[TIAB:~1] OR "literature searches"[TIAB:~1] OR "narrative review"[TIAB:~1] OR "narrative reviews"[TIAB:~1] OR "qualitative review"[TIAB:~1] OR "qualitative reviews"[TIAB:~1] OR "quantitative review"[TIAB] OR "quantitative reviews"[TIAB])) OR "comprehensive review"[TIAB] OR "comprehensive reviews"[TIAB] OR "comprehensive search"[TIAB] OR "comprehensive searches"[TIAB] OR "critical review"[TIAB] OR "critical reviews" [TIAB] OR (("electronic database"[TIAB:~1] OR "electronic databases"[TIAB:~1] OR "databases searched"[TIAB:~3]) AND (eligibility[tiab] OR excluded[tiab] OR exclusion[tiab] OR included[tiab] OR inclusion[tiab])) OR "evidence assessment"[TIAB] OR "evidence review"[TIAB] OR "exploratory review"[TIAB] OR "framework synthesis"[TIAB] OR "Integrated review"[TIAB] OR "integrated reviews"[TIAB] OR "integrative review"[TIAB:~1] OR "integrative reviews"[TIAB:~1] OR "mapping review"[TIAB:~1] OR "meta-review"[TIAB:~1] OR "meta-synthesis"[TIAB:~1] OR "methodology review"[TIAB:~1] OR ("mixed methods"[TIAB:~0] AND "methods review"[TIAB:~1]) OR ("mixed methods"[TIAB:~0] AND "methods synthesis"[TIAB:~1]) OR "overview reviews"[TIAB:~4] OR ("PRISMA"[TIAB] AND "preferred"[TIAB]) OR "PRISMA-P"[TIAB:~0] OR "prognostic review"[TIAB:~1] OR "psychometric review"[TIAB:~1] OR ("rapid evidence"[TIAB:~0] AND "evidence assessment"[TIAB:~0]) OR "rapid realist"[TIAB:~0] OR "rapid review"[TIAB:~1] OR "rapid reviews"[TIAB:~1] OR "realist review"[TIAB:~1] OR "review of reviews"[TIAB:~1] OR "scoping review"[TIAB:~1] OR "scoping reviews"[TIAB:~1] OR "scoping study"[TIAB:~1] OR ("state art "[TIAB:~2] AND "art review"[TIAB:~1]) OR "systematic evidence map"[TIAB] OR "systematic mapping"[TIAB:~1] OR "systematic literature"[TIAB:~1] OR "systematic Medline"[TIAB:~2] OR "systematic PubMed"[TIAB:~2] OR "systematic review"[TIAB:~1] OR "systematic reviews"[TIAB:~1] OR "systematic search"[TIAB:~1] OR "systematic searches"[TIAB:~1] OR "systematical review"[TIAB:~1] OR "systematical reviews"[TIAB:~1] OR "systematically identified"[TIAB:~1] OR "systematically review"[TIAB:~1] OR "systematically reviewed"[TIAB:~1] OR "umbrella review"[TIAB:~1] OR "umbrella reviews"[TIAB:~1] OR "Cochrane Database Syst Rev"[ta] OR "evid rep technol assess full rep"[Journal] OR "evid rep technol assess summ"[Journal])) |
