## Supplementary material for "The Effect of Psychotherapy in the Management of Obesity: An umbrella review": Table 1. Main findings provided by the selected articles

| Título | Autores | Año | Tipo de estudio | Revista | Referencia | Tipo de psicoterapias evaluadas | Objetivos | Resultados primarios | Resultados secundarios | Link | Extra data |
|---|---|---|---|---|---|---|---|---|---|---|---|
| Family-based models for childhood-obesity intervention: A systematic review of randomized controlled trials | P. Sung-Chan, Y. W. Sung, X. Zhao and R. C. Brownson | 2013 | Systematic review | obesity reviews | doi: 10.1111 /obr.12000 | Family based interventions that include family therapy | This review aimed to examine the methodological rigour<br><br>and treatment effectiveness of family-based interventions according to interven-tion types and theoretical orientations. | Rigour: Based on the results of MQRS, the overall methodological rigour of the 15 RCTs was satisfactory. The MQRS ranged from six to 12 points, with a median score of 8 and an average score of 7.93. Ten studies (67%) received a score of eight or above (out of 14), while the remaining five studies (33%) scored an average of 6.2 (see Fig. 2). These findings suggest that 10 out of 15 RCTs had well aligned their research questions with appropriate and rigourous research methods.<br><br>Interventions: the greatest effect was achieved with the family based lifestyle intervention, lifestyle intervention plus parent education and family theraphy | | https://onlinelibrary.wiley.com/doi/abs/10.1111/obr.12000 | 15 RCT<br>Heterogenity: was considerable, making difficult to analyze the results, but there was no specific data |
| Comparative Efectiveness of Cognitive Behavioral Therapy and Behavioral Therapy in Obesity: A Systematic Review and Network Meta-Analysis | Katja Kurnik Mesarič, Jernej Pajek, Bernarda Logar Zakrajšek, Špela Bogataj & Jana Kodrič | 2023 | systematic review and meta-analysis | Nature | https://doi.org/10.1038/s41598-023-40141-5 | cognitive behavioral therapy (CBT) | The aim of this systematic review and meta-analysis was to examine the contribution of cognitive behavioral therapy (CBT) to the implementation of lifestyle changes, considering health-related and behavioral outcomes. | In most of the studies included in this review (where weight loss was the goal of the intervention), the interven-tions were found to have had at least some efect on weight loss17–19,22. In two of the studies, the intervention had<br><br>no efect on weight loss15,16. Tree of the included studies reported data for weight change in BMI immediately afer the weight loss intervention. | | https://www.nature.com/articles/s41598-023-40141-5 | 33 RCT<br>Posible metodological heterogenity |
| Weight-loss interventions for improving emotional eating among adults with high body mass index: A systematic review with meta-analysis and meta-regression | Han Shi Jocelyn Chew, Siew Tiang Lau, Ying Lau | 2023 | systematic review with meta-analysis and meta-regression | Eur Eat Disorders | Eur Eat Disorders Rev. 2022;30:304–327. | Cognitive Behaviour Therapy (CBT) | To evaluate the effectiveness of weight-loss interventions on emotional eating among adults with high body mass index (BMI). | Results suggested a small-to-medium interventional ef-fect on emotional eating factor post-intervention.<br>Results suggested<br><br>a small-to-medium interventional effect on uncontrolled eating post-intervention<br>Results suggested a small-to-medium interventional effect on the cognitive restraint factor directly post-intervention<br>Results suggested no significant interventional effect on binge-eating behaviour post-intervention | Results suggested no significant interventional effect on weight measured in BMI directly post-intervention<br><br>Results suggested no significant effect of emotional eating interventions on the participants' weight measured in kg directly post-intervention | https://onlinelibrary.wiley.com/doi/10.1002/erv.2906 | 31 RCT<br>Age, male proportion, baseline BMI, attrition rate and intervention length were not significant moderators of the heterogeneity between studies. |
