## Supplementary material for "The Effect of Psychotherapy in the Management of Obesity: An umbrella review": Figure 1: Flow diagram of the identification, screening, and selection of studies for this Umbrella review.

**PRISMA 2020 flow diagram for new systematic reviews which included searches of databases and registers only**

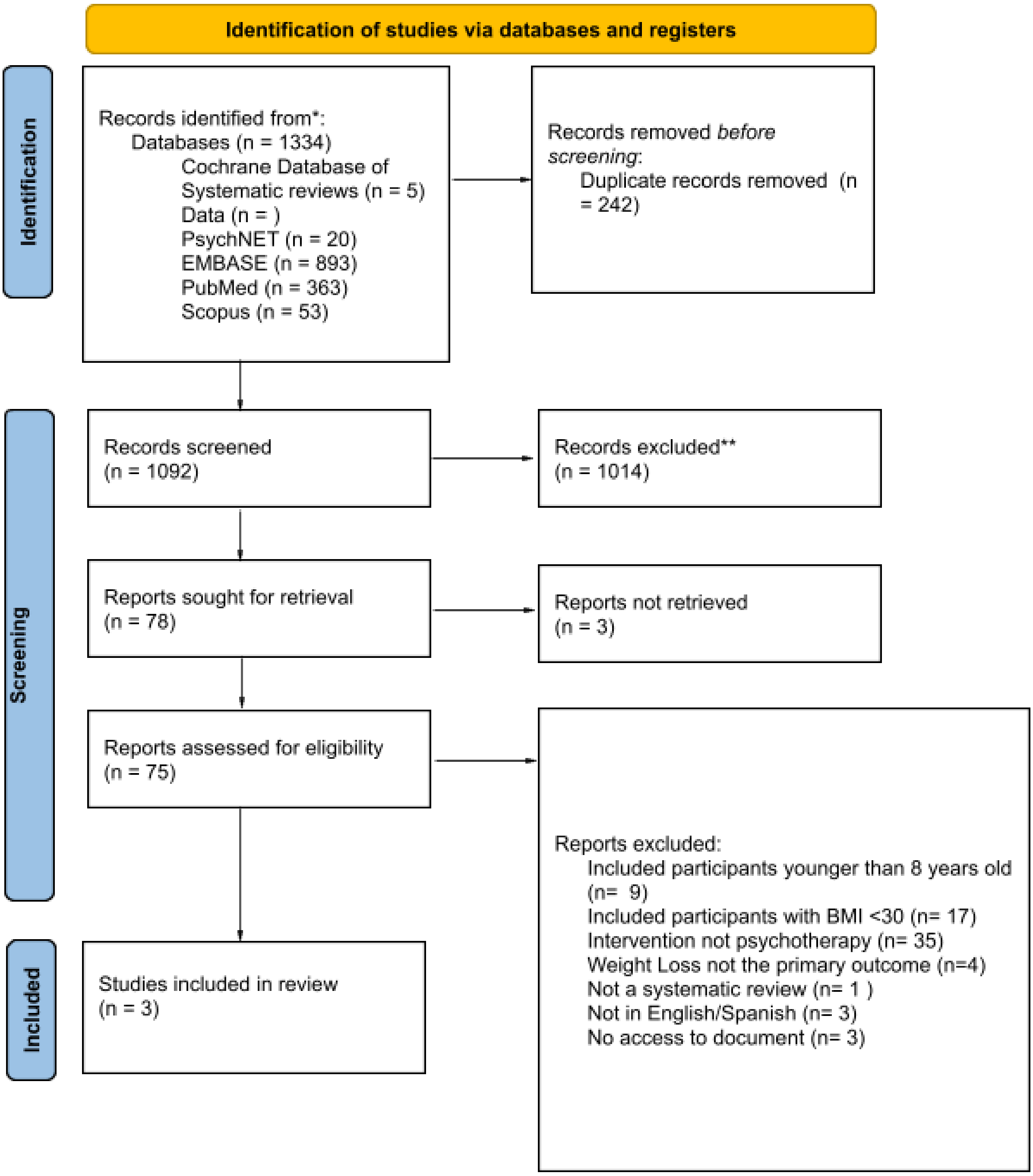


Figure 1: Flow diagram of the identification, screening, and selection of studies for this Umbrella review.

The study selection process was conducted in several data bases, indexed terms were used to aid the searches. A total of 1334 articles were retrieved. Results were imported to Rayyan QCI software, after excluding duplicates both manually and utilizing the automated duplicate identification tool,1092 articles were left. Then, articles were screened by title and abstract, taking into consideration the selection criteria established, only 78 reports met these criteria and were taken under full-text revision, 3 reports could not be accessed to, while 72 articles were excluded because they did not meet the selection criteria. Articles excluded were classified according to the following categories: 1. Included participants younger than 8 years old (9) 2. Included participants with BMI <30 (17) 3. Intervention not psychotherapy (35) 4. Weight loss not the primary outcome (4) 5. Not a systematic review (1) 6. Not in English/Spanish (3) and 7. no access to document (3). Additionally, there were several articles which fell into more than one of these categories, for instance, there were n articles that had both wrong population and wrong intervention.

*From:* Page MJ, McKenzie JE, Bossuyt PM, Boutron I, Hoffmann TC, Mulrow CD, et al. The PRISMA 2020 statement: an updated guideline for reporting systematic reviews. BMJ 2021;372:n71. doi: 10.1136/bmj.n71
